# Genetic associations with severe COVID-19

**DOI:** 10.1101/2021.03.29.21254509

**Authors:** Nicholas M. Murphy, Gillian S. Dite, Richard Allman

## Abstract

Identification of host genetic factors that predispose individuals to severe COVID-19 is important, not only for understanding the disease and guiding the development of treatments, but also for risk prediction when combined to form a polygenic risk score (PRS). Using population controls, Pairo-Castineira et al. identified 12 SNPs (a panel of 8 SNPs and a panel of 6 SNPs, with two SNPs in both panels) associated with severe COVID-19. Using controls with asymptomatic or mild COVID-19, we were able to replicate the association with severe COVID-19 for only three of their SNPs and found marginal evidence for an association for one other. When combined as an 8-SNP PRS and a 6-SNP PRS, we found no evidence of association with severe COVID-19. The difference in our results and the results of Pairo-Castineira et al. might be the choice of controls: population controls vs controls with asymptomatic or mild COVID-19.

## Paper

Pairo-Castineira et al.^1^ identified 12 single nucleotide polymorphisms (SNPs) that were associated with critical illness in COVID-19, in particular a panel of 8 SNPs (associated with risk of intensive care admission) from independent genome-wide significant regions and a panel of 6 SNPs (associated with hospitalization) from a meta-analysis of overlapping SNPs between GenOMICC, Host Genetics Initiative and 23andMe studies (two of these SNPs were also in the panel of 8 SNPs). These SNP panels were identified and validated using population controls.

Using the UK Biobank,^2-4^ we evaluated the ability of the two SNP panels to predict severe COVID-19 – should a person become infected – when combined as PRSs. We downloaded the UK Biobank COVID-19 results file on 8 January 2021 and identified 8672 active participants with a positive SARS-CoV-2 test result, 8374 of whom had SNP data available. As we did previously,^5^ we used source of test result as a proxy for disease severity, with 2288 (27.3%) participant results coming from an inpatient setting (cases) and 6086 (72.7%) participant results coming from an outpatient setting (controls).

We used the estimates of the odds ratio (OR) per effect allele provided in Pairo-Castineira et al.^1^ to construct the 8-SNP and 6-SNP PRS (the meta-analysis ORs for the 6-SNP PRS). For the SNPs in their Table 1 and the two overlapping SNPs from their Table 2, we used the GenoMICC risk allele frequencies provided, and for the remaining SNPs in their Table 2, we used allele frequencies from SNPnexus.^6^ For the construction of the PRS,^7^ we assumed independent and additive risks on the log OR scale. For each SNP, we calculated the unscaled population average risk (µ) as:

**Table 1.**
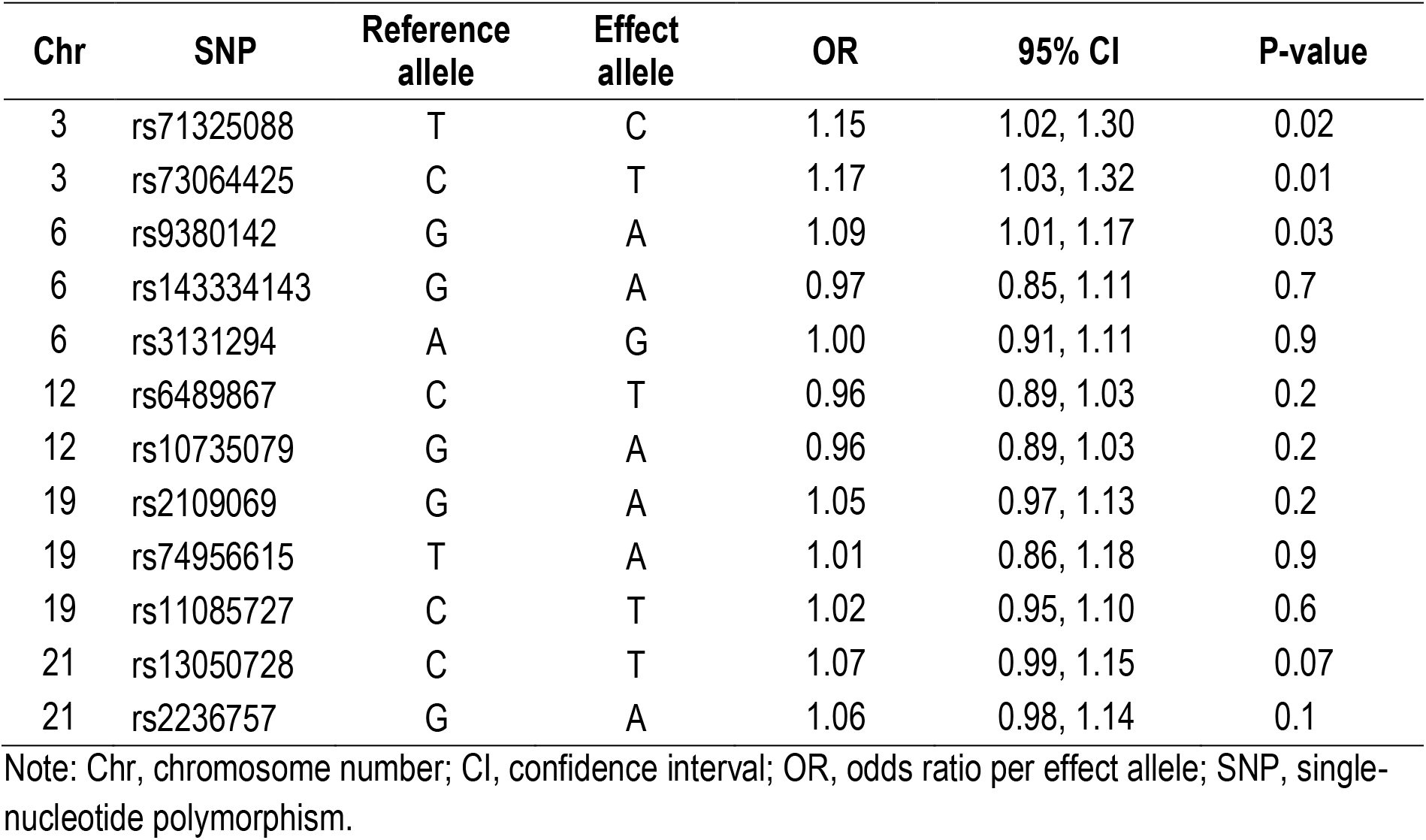
Unadjusted odds ratios per effect allele for the 12 SNPs.

| Chr | SNP | Reference allele | Effect allele | OR | 95% CI | P-value |
| --- | --- | --- | --- | --- | --- | --- |
| 3 | rs71325088 | T | C | 1.15 | 1.02, 1.30 | 0.02 |
| 3 | rs73064425 | C | T | 1.17 | 1.03, 1.32 | 0.01 |
| 6 | rs9380142 | G | A | 1.09 | 1.01, 1.17 | 0.03 |
| 6 | rs143334143 | G | A | 0.97 | 0.85, 1.11 | 0.7 |
| 6 | rs3131294 | A | G | 1.00 | 0.91, 1.11 | 0.9 |
| 12 | rs6489867 | C | T | 0.96 | 0.89, 1.03 | 0.2 |
| 12 | rs10735079 | G | A | 0.96 | 0.89, 1.03 | 0.2 |
| 19 | rs2109069 | G | A | 1.05 | 0.97, 1.13 | 0.2 |
| 19 | rs74956615 | T | A | 1.01 | 0.86, 1.18 | 0.9 |
| 19 | rs11085727 | C | T | 1.02 | 0.95, 1.10 | 0.6 |
| 21 | rs13050728 | C | T | 1.07 | 0.99, 1.15 | 0.07 |
| 21 | rs2236757 | G | A | 1.06 | 0.98, 1.14 | 0.1 |

Note: Chr, chromosome number; CI, confidence interval; OR, odds ratio per effect allele; SNP, single-nucleotide polymorphism.

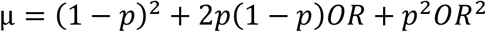

Next, for each SNP, adjusted risks (with a population average risk equal to 1) were calculated as:

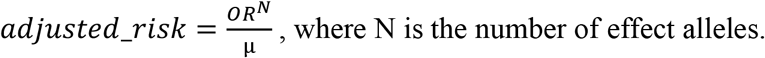

The PRS was then calculated as the product of the adjusted risk values for each of the\ SNPs.

We first used logistic regression to estimate the unadjusted per allele effects for each SNP (see Table 1). Of the 12 SNPs, three (rs71325088, rs73064425, and rs9380142) were associated with severe disease and one (rs13050728) was marginally statistically significant using a nominal statistical significance of P<0.05. These SNPs all had smaller effect sizes in our analysis compared with those reported in Pairo-Castineira et al.^1^ (rs71325088, OR=1.15 vs 1.9, respectively; rs73064425, OR=1.17 vs 2.1; rs9380142, OR=1.09 vs 1.3; and rs13050728, OR=1.07 vs 1.2).

We then used logistic regression to estimate the OR per quintile of risk in the controls for the two PRS. While marginally statistically significant, these showed only a weak association with a 3% increased risk of severe COVID-19 per quintile of risk (for both: OR=1.03; 95% confidence interval [CI]=1.00, 1.07; P=0.05). The two PRS showed almost no ability to discriminate cases from controls, with an area under the receiver operating characteristic curve (AUC) for the 8-SNP PRS of 0.514 (95% CI=0.500, 0.528) and an AUC for the 6-SNP PRS of 0.512 (95% CI=0.498, 0.526).

Using population controls rather than people who are SARS-CoV-2 positive but are asymptomatic or have mild disease may have done more than bias associations towards the null, as suggested by Pairo-Castineira et al.^1^ If the associations seen using population controls were biased towards the null, we would expect the effects of loci identified with population controls to be stronger when using asymptomatic or mild SARS-CoV-2 positive controls.

This was not evident in our analyses nor was it evident in the COVID-19 Host Genetics Initiative^8,9^ Release 5 Manhattan plots (for the *Hospitalized COVID-19 vs not-hospitalized COVID-19* and the *Hospitalized COVID-19 vs population controls* meta-analyses), where other than the 3p21.31 locus, the regions of association differ considerably. In the *Hospitalized COVID-19 vs population controls* meta-analysis, the only region of association is at 3p21.31, while for the *Hospitalized COVID-19 vs population controls* meta-analysis, 3p21.31 was a clear region of association and there were six other regions of association (1q21.3, 9q34.2, 12q24.13, 17q21.33, 19p13.3, and 21q21.11; see Figure 1A and B).

**Figure 1.**
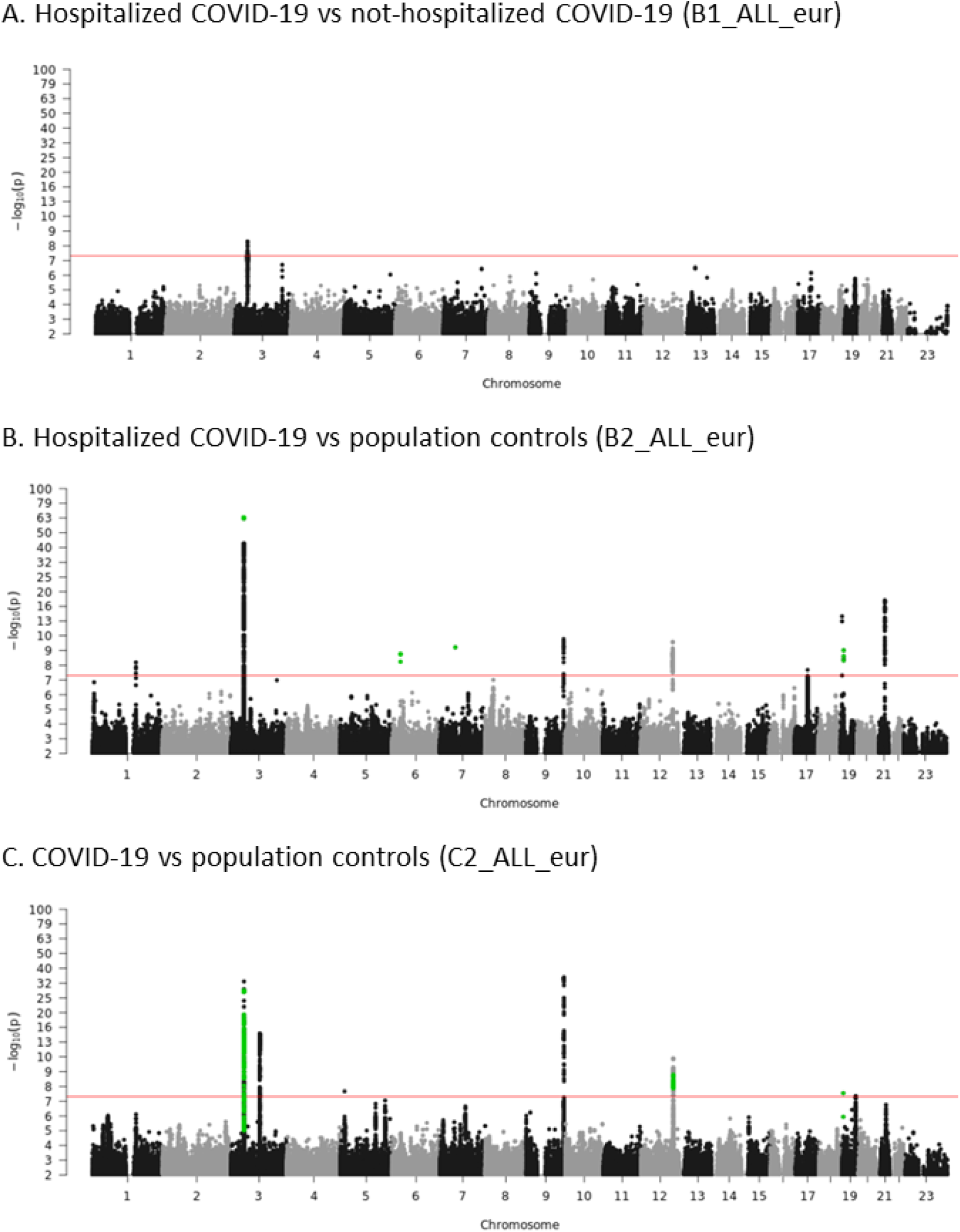
Manhattan plots from the COVID-19 Host Genetics Initiative Release 5 meta-analyses of (A) *Hospitalized COVID-19 vs not-hospitalized COVID-19, Hospitalized COVID-19 vs population controls*, and (C) *COVID-19 vs population controls*.

Interestingly, three of these regions (3p21.31, 9q34.2, and 12q24.13) overlapped with the Manhattan plot for all COVID-19 (i.e. infection): the *COVID-19 vs population controls* meta-analysis (see Figure 1B and C). The 3p21.31 locus has been identified previously in studies that used population controls,^10,11^ and rs71325088 and rs73064425 SNP identified by Pairo-Castineira et al.^1^ are both in 3p21.31. We also see an association for those two SNPs, albeit smaller in magnitude, using SARS-CoV-2 positive controls.

The use of population controls might mean that Pairo-Castineira et al.^1^ have identified SNPs associated with becoming infected with SARS-CoV-2 rather than SNPs associated with severe COVID-19, a limitation that has been identified by other authors using population controls.^12^ Researchers who use the results of large studies with population controls to inform their investigation of severe COVID-19 may therefore be wasting their time and resources. We urge care in the interpretation of results from large-scale studies that use population controls and invite discussion on this issue.

## Data Availability

The data underlying this article was provided by the UK Biobank and we do not have permission to share the data. Researchers wishing to access the data used in this study can apply directly to the UK Biobank at https://www.ukbiobank.ac.uk/register-apply/. Stata 16.1 code for the analysis is available from the corresponding author on request.

## Ethics approval

The UK Biobank has Research Tissue Bank approval (REC #11/NW/0382) that covers analysis of data by approved researchers. All participants provided written informed consent to the UK Biobank before data collection began. This research has been conducted using the UK Biobank resource under Application Number 47401.

## Author contributions

All authors were involved in the conceptual development of this study. NMM and RA examined the Host Genetics Initiative meta-analysis results. GSD undertook data management and conducted the statistical analyses. All authors contributed to writing the first draft and reviewed and edited the manuscript. All authors have approved the final manuscript.

